# Antibody and T cell responses to a single dose of the AZD1222/Covishield vaccine in previously SARS-CoV-2 infected and naïve health care workers in Sri Lanka

**DOI:** 10.1101/2021.04.09.21255194

**Authors:** Chandima Jeewandara, Achala Kamaladasa, Pradeep Darshana Pushpakumara, Deshni Jayathilaka, Inoka Sepali Abayrathna, Saubhagya Danasekara, Dinuka Guruge, Thushali Ranasinghe, Shashika Dayarathne, Thilagaraj Pathmanathan, Gayasha Somathilaka, Deshan Madhusanka, Shyrar Tanussiya, Tibutius TPJ, Heshan Kuruppu, Ayesha Wijesinghe, Nimasha Thashmi, Dushantha Milroy, Achini Nandasena, Nilanka Sanjeewani, Ruwan Wijayamuni, Sudath Samaraweera, Lisa Schimanski, T.K. Tan, Tao Dong, Graham S. Ogg, Alain Townsend, Gathsaurie Neelika Malavige

**Author notes:** Correspondence should be addressed to: Prof. Neelika Malavige DPhil (Oxon), FRCP (Lond), FRCPath (UK), AICBU, Department of Immunology and Molecular Medicine, Faculty of Medical Sciences, University of Sri Jayawardanapura, Sri Lanka. contributed equally (Chandima Jeewandara, Achala Kamaladasa, Pradeep Darshana Pushpakumara, Deshni Jayathilaka).

## Abstract

**Background:** In order to determine the immunogenicity of a single dose of the AZD1222/Covishield vaccine in a real-world situation, we assessed the immunogenicity, in a large cohort of health care workers in Sri Lanka.

**Methods:** SARS-CoV-2 antibodies was carried out in 607 naïve and 26 previously infected health care workers (HCWs) 28 to 32 days following a single dose of the vaccine. Haemagglutination test (HAT) for antibodies to the receptor binding domain (RBD) of the wild type virus, B.1.1.7, B.1.351 and the surrogate neutralization assay (sVNT) was carried out in 69 naïve and 26 previously infected individuals. Spike protein (pools S1 and S2) specific T cell responses were measured by *ex vivo* ELISpot IFNγ assays in 76 individuals.

**Results:** 92.9% of previously naive HCWs seroconverted to a single dose of the vaccine, irrespective of age and gender; and ACE2 blocking antibodies were detected in 67/69 (97.1%) previously naïve vaccine recipients. Although high levels of antibodies were found to the RBD of the wild type virus, the titres for B.1.1.7 and B.1.351 were lower in previously naïve HCWs. *Ex vivo* T cell responses were observed to S1 in 63.9% HCWs and S2 in 31.9%. The ACE2 blocking titres measured by the sVNT significantly increased (p<0.0001) from a median of 54.1 to 97.9 % of inhibition, in previously infected HCWs and antibodies to the RBD for the variants B.1.1.7 and B.1.351 also significantly increased.

**Discussion:** a single dose of the AZD1222/Covishield vaccine was shown to be highly immunogenic in previously naïve individuals inducing antibody levels greater than following natural infection. In infected individuals, a single dose induced very high levels of ACE2 blocking antibodies and antibodies to RBDs of SARS-CoV-2 variants of concern.

**Funding:** We are grateful to the World Health Organization, UK Medical Research Council and the Foreign and Commonwealth Office.

## Introduction

The first cases of COVID-19 due to infection with the SARS-CoV-2 infection were reported in December 2019, from Wuhan in the Hubei province in China ^1^. However, within one year, not only were several types of vaccines for COVID-19 developed, but they were used in mass immunization campaigns in many parts of the world, after successful completion of phase 3 trials ^2-4^. The mRNA COVID-19 vaccines Pfizer-BioNTech received emergency use authorization on 11^th^ December 2020 in and the Moderna on the 18^th^ of December USA, while the UK MHRA approved the AstraZeneca vaccine on the 30^th^ of December 2020 ^2,4^. The mass scale immunization campaigns that were initiated in December and early January 2021, have already shown to be effective by significantly reducing deaths, severe disease and hospitalizations in groups that received these vaccines ^5,6^.

While most of the vaccines for prevention of COVID-19 are two dose vaccines, some vaccines such as the Johnson and Johnson adenoviral vector vaccine comprise a single dose, reporting an efficacy rate of 66% against symptomatic infection and 85% efficacy against severe disease ^7^. Although the efficacy of a single dose administration of the other WHO approved vaccines has not been evaluated in large clinical trials, in some countries, in order to administer the first dose to a larger population, the second dose was delayed for up to 12 weeks ^8^. A single dose of both the BNT162b2 (Pfizer BioNTech) vaccine and the AZD1222 (Astrazeneca) adenoviral vector vaccine was found to significantly reduce hospitalizations due to COVID-19, 28 to 34 days since administration of the first dose ^9^. It was recently shown that a single dose of the BNT162b2 (Pfizer BioNTech) vaccine induced T cell and antibody responses that were comparable to those who were naturally infected with the SARS-CoV-2, several weeks or months following infection ^10^. Although these data suggest that in a pandemic situation, where most countries have a shortage of vaccines, administering a single dose of a two-dose vaccine, does indeed offer substantial protection, there has been criticism that such an approach would give rise to the emergence of variants, due to a suboptimum immune response in those who only receive a single dose of a vaccine ^8,11^. Those especially with haematological malignancies were shown to have a suboptimal immune response to a single dose of the BNT162b2 (Pfizer BioNTech), which leave them vulnerable to infection with the SARS-CoV-2 and for potential emergence of new variants ^12^. However, some countries such as Canada have decided to delay the second dose for 16 weeks, despite these concerns ^13^.

There have been many variants of concern which are due to mutations in the spike protein of the virus, which either increase disease transmission, evade detection by currently available diagnostics or the mutations are in major sites where neutralizing antibodies bind to, and therefore, they have a potential to affect vaccine efficacy^14^. The B.1.1.7 variant, which was initially detected in the UK, has shown to associate with higher transmissibility and higher mortality rates^14,15^. Although AZD1222 and BNT162b2 (Pfizer BioNTech) have shown a slightly reduced neutralization activity against B.1.1.7, it did not have a significant impact on vaccine efficacy ^16,17^. However, the E484K mutation present in both the B.1.351 variant and P.1 variant have shown to significantly affect the neutralizing ability of the antibodies generated by most vaccines ^16-18^. Since most of the COVID-19 vaccines underwent clinical trials, when these particular variants were not dominant, it would be important to determine the immune responses generated by these vaccines in neutralizing these variants of concern.

Although many developed countries such as the UK, Europe and USA have administered one dose of a COVID-19 vaccine to over 15% of their population by 1^st^ of April 2021, many South Asian and South East Asian countries have administered one dose for <5%, while some African and Asian countries have immunized <1% ^19^. Therefore, many countries in the world would have a partially immunized population, with a single vaccine dose administered. Furthermore, due to recent concerns regarding possible side effects such as cerebral venous thrombosis and thrombocytopenia, in relation to the AZD1222 vaccine^20^, many individuals in some countries appear to be hesitant to obtain the second dose. In order to determine the immunogenicity of a single dose of the AZD1222/Covishield vaccine in a real time situation, we assessed the immunogenicity (antibody and T cell responses), in a large cohort of health care workers in Sri Lanka, who received the AZD1222/Covishield vaccine during late January/early February and we also assessed the immune responses generated by these vaccines against the variants of concern (B.1.1.7 and B.1.351).

## Results

The demographic characteristics and previous infection status of the 633 health care workers (HCWs) is shown in table 1. 26/633 (4.26%) of individuals had past infection with the SARSCoV-2. The median age of the HCWs was 41 years (range 21 to 81 years). 367 (57.9%) were females and 50 (7.9%) had at least one comorbid condition (hypertension, diabetes or chronic kidney disease). The overall seroconversion after a single dose was 588 (92.9%). The seropositivity of these individuals between 28 to 32 days since obtaining the first dose of the vaccine is shown in table 1. The seroconversion rates were highest in the 40 to 49 age group, whereas the seroconversion rates were lower in those >60 years of age 81.6%. Seroconversion rates were equal among males (244, 92.8%) and females (343, 93.4%). There was no difference in the SARS-CoV-2 total antibody levels between males (median 7.1, IQR 3.57 to 11.47 antibody index), compared to females (median 7.7, IQR 4.1 to 11.81). Of the 50 individuals who had comorbidities, 48 (96%) seroconverted.

**Table 1:**
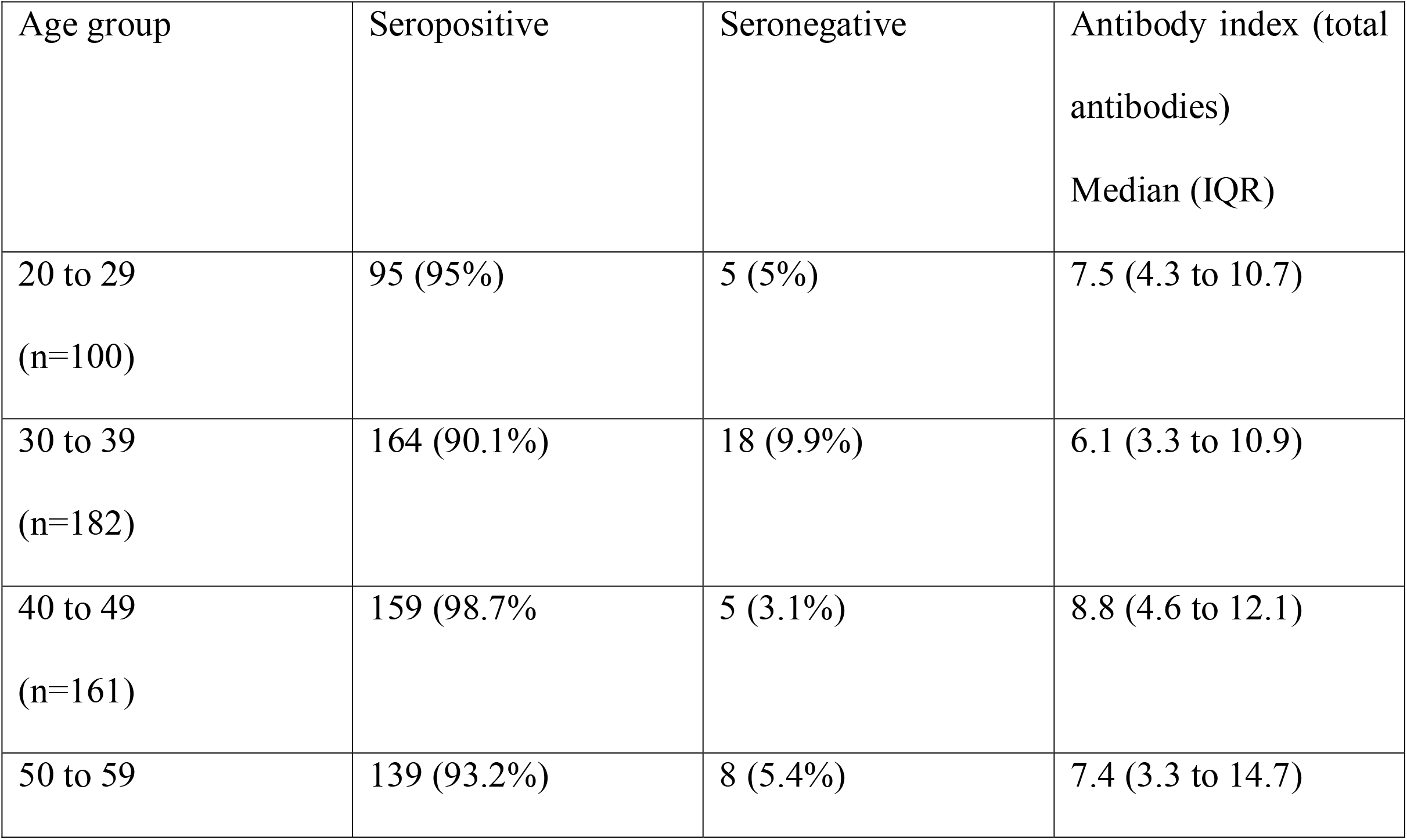
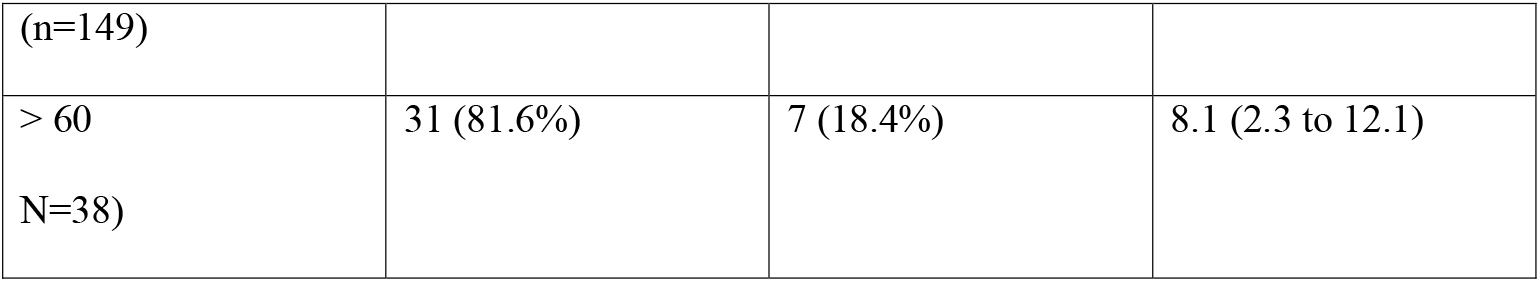
Seropositivity rates of a single dose of the ChadOx1 between 28 to 32 days in HCWs.

### Antibody titres in naïve individuals and those who were immune to the SARS-CoV-2

The antibody index is an indirect measurement of the antibody levels of this SARS-CoV-2 total antibody assay. The median antibody titres were lowest in the 30- to 39-year-old age group (median 6.1, IQR= 3.3 to 10.9 index value) compared to other age groups. The levels in >60 age group showed a median of 8.1 (IQR=2.3 to 12.13 index value) and this difference was statistically significant (p=0.03) (Figure 1 A). The antibody index values of those who had past COVID-19 at the time of recruitment was a median of 8.9 (IQR 2.6 to 11.9), which significantly (p<0.0001) rose to a median of 13.1 (IQR12.5 to 14.0) between 28 to 32 days following a single dose of the vaccine (Figure 1B).

**Figure 1:**
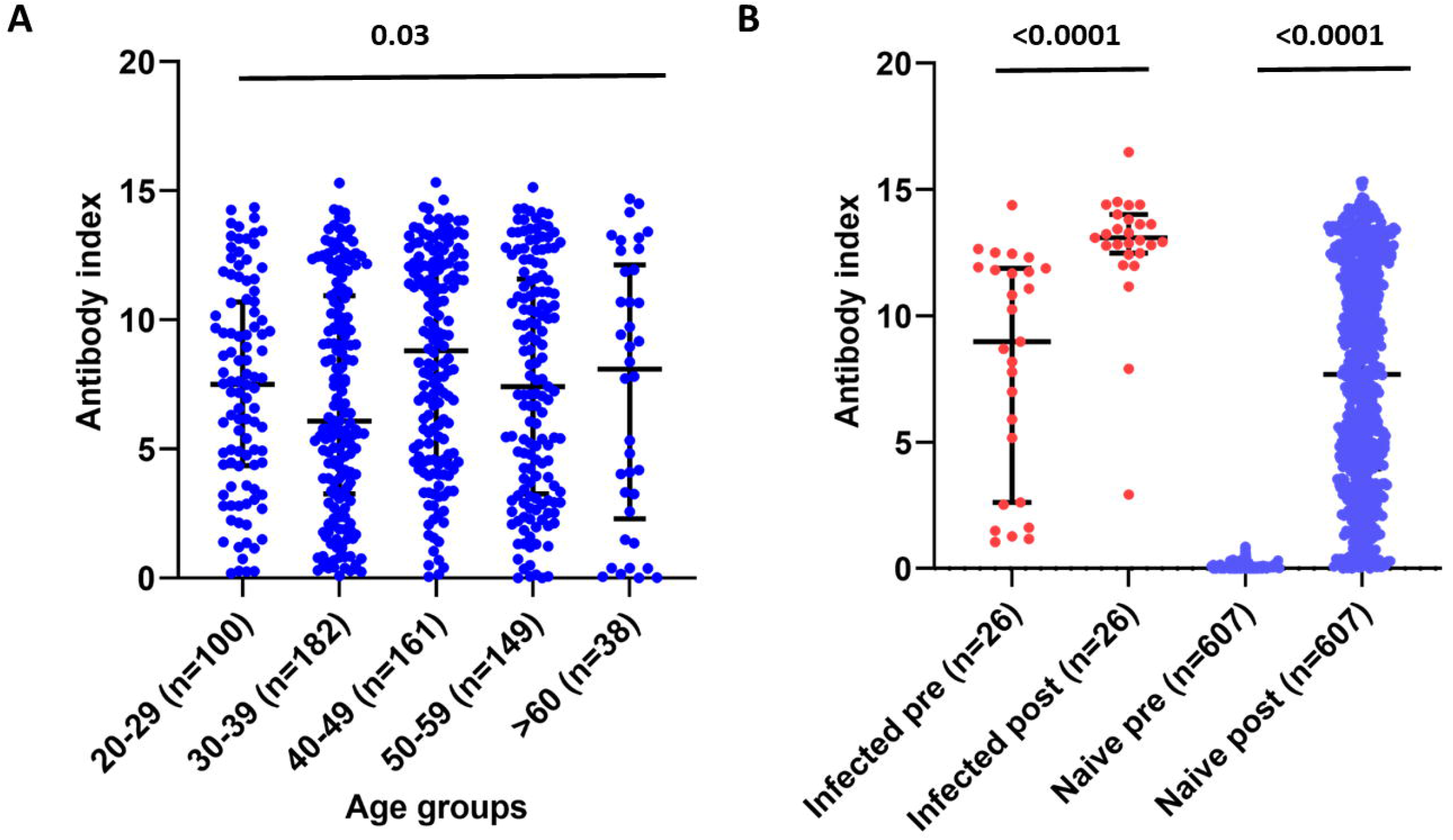
SARS-CoV-2 specific total antibody levels in vaccinated individuals. SARS-CoV-2 total antibody levels (antibody index) in those in different age groups (A), and the total antibody levels in those who had previous infection at baseline and 28 to 32 days after a single dose, and in SARS-CoV-2 uninfected individuals at baseline and 28 to 32 days after a single dose (B) were measured using the WANTAI SARS-CoV-2 Ab ELISA assay.

### Antibodies to the Receptor Binding Domain of the spike protein, measured by the Haemagglutination test

Haemagglutination test (HAT) to measures antibodies to the RBD where the RBD of the virus is linked to a nanobody IH4, specific for a conserved epitope within glycophorin A on red blood cells (RBCs)^21^. We have confirmed that this assay is negative in >99% of individuals prior to infection with SARS-CoV-2. We then used the assay to measure antibody titres to the RBD of the SARS-CoV-2 wild type (WT) virus, B.1.1.7 variant and the B.1.351 variant in 69 individuals who were SARS-CoV-2 seronegative, and 26 individuals who had been infected with the virus. The median post-vaccination HAT titres of those who were seronegative at baseline was 1:40 to the WT, 1:20 to B.1.1.7 and 0 to B.1.351, 28 to 32 days following a single dose of the vaccine (Figure 2A). Following a single dose of the vaccine, those who had past COVID-19 had significantly higher HAT titres to the WT (p<0.0001), B.1.1.7 (p<0.0001) and the B.1.351 (p<0.0001) (Figure 2A). While the SARS-CoV-2 naïve individuals had significantly less (p<0.0001) HAT titre to the B.1.1.7 compared to the WT following immunization, there was no significant differences in the HAT titres to WT and B.1.1.7 (p=0.21) in those who were seropositive for SARS-CoV-2, at the baseline. Both groups of individuals who were seronegative and seropositive at baseline, had significantly less (p<0.0001) HAT titres to the B.1.351, compared to the WT and B.1.1.7. Following a single dose of the vaccine, those who were seropositive at baseline, had a significant increase in the HAT titres for WT (p=0.005), B.1.1.7 (p<0.0001) and B.1.351 (p=0.0004) (Figure 2B).

**Figure 2:**
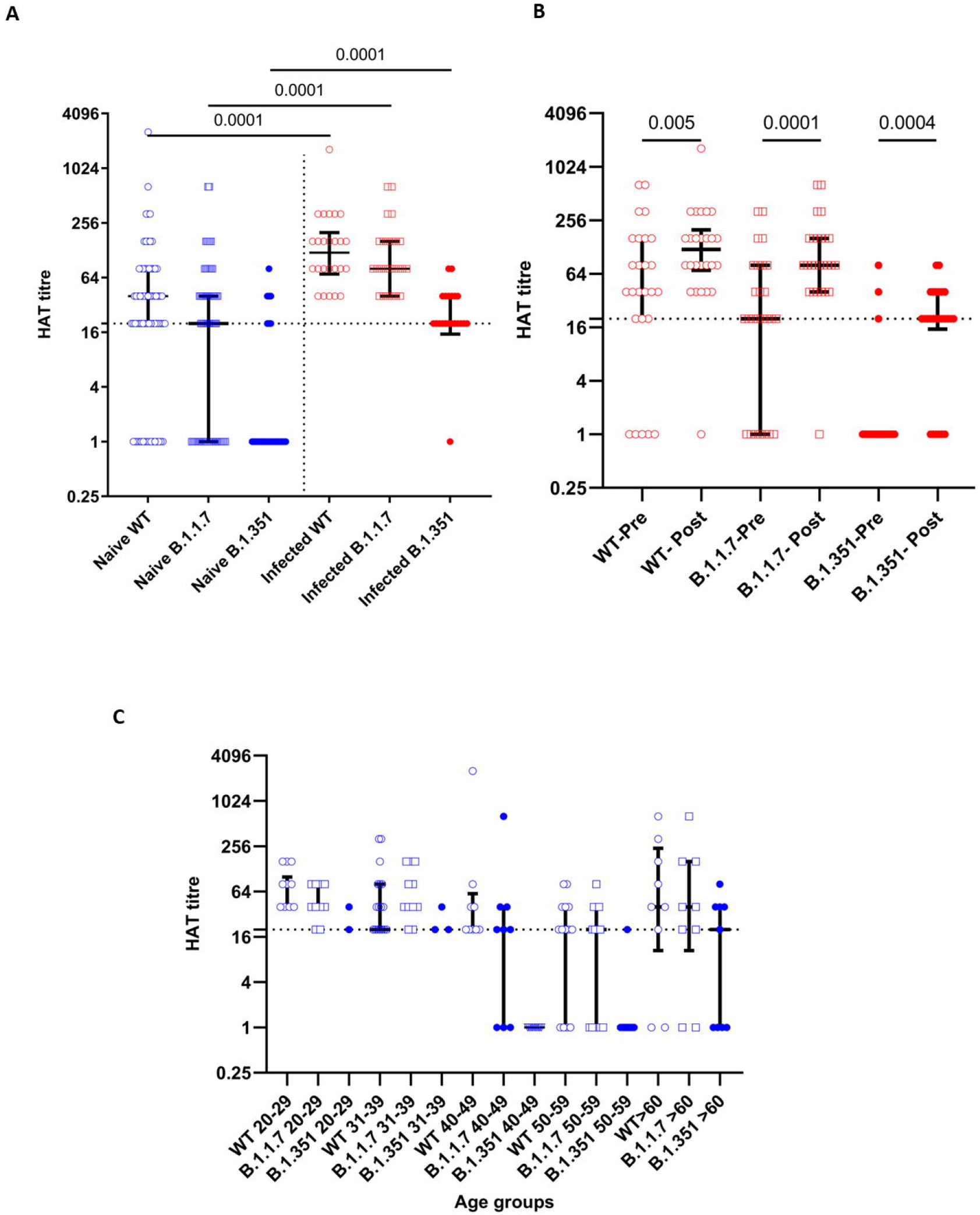
Haemagglutination test to detect antibodies to RBD of the wild type (WT), B.1.1.7 and B.1.351 in patients who were naïve and previously infected following a single dose of the AZD1222. The HAT titres for the WT, B.1.1.7 and B.1.351 were measured in naïve individuals (blue) and previously infected individuals (red) 28-32 days following the vaccine (A). The HAT titres were measured in previously infected individuals at the baseline and following vaccination for the WT, B.1.1.7 and B.1.351 (B). The HAT titres were measured following a single dose in previously naïve individuals in different age groups (C). The black dotted line indicates the positive cut of for the HAT

A HAT titre of 1:20 was considered as positive for the presence RBD-specific antibodies. 54/69 (78.2%) of individuals who were seronegative, had a positive RBD antibodies following a single dose of the vaccine. 45 (65.2%) had positive responses to the RBD of B.1.1.7 and 11 (15.9%) had responses to the RBD of B.1.351. At the baseline 21/26 (80.76%) who were known to be infected previously with the SARS-CoV-2, had antibodies to the RBD of the WT virus. 19/26 (73%) had antibodies to RBD of B.1.1.7 and only 3/26 (11.5%) had antibodies to RBD of B.1.351. However, following a single dose of the vaccine, 25/26 (96.1%) developed antibodies to RBD of the WT, 25/26 (96.1%) to the RBD of B.1.1.7 and 20/26 (76.9%) to the RBD of B.1.351. There was no significant difference between HAT titres to the RBD of the WT, B.1.1.7 (Figure 2C). However, there was a significance difference in the titres for the B.1.351 (p=0.006), as those >60 years of age, had higher titres than some age groups (40 to 49 age group). This is possibly due to lower sample size in certain age groups. For instance, in the 40 to 49 age group (n=9), no one had any antibodies to the RBD of B.1.351, whereas in the >60 age group (n=9), 5 had IgG antibodies.

### Surrogate neutralization assay to assess ACE2 blocking antibodies following a single dose of the AZD1222

Due to the absence of facilities to carry out live virus assays to detect the presence of neutralizing antibodies (NAbs), we used a surrogate assay, which measured ACE2 blocking antibodies has been shown to correlate with the NAbs specific for the SARS-CoV-2^22^. The sVNT titres (percentage of inhibition of ACE2 binding) significantly increased 28 to 32 days post vaccination in previously naïve individuals (p<0.0001) and in previously infected individuals (p<0.0001) (Figure 3A). However, those who were previously infected with the SARS-CoV-2 (median 97. 99, IQR 89.65 to 99.27 % of inhibition) had significantly higher levels (p<0.0001) than those who were naïve (median 69.42, IQR 54.09 to 81.54 % of inhibition). Only 2/69 (2.9%) individuals who were previously naïve failed to develop the level of 25% inhibition (regarded as “positive”) following a single dose of the vaccine. Of those who were seropositive at recruitment, 6/26 (23.1%) were negative for the presence of ACE2 blocking antibodies by sVNT (<25% of inhibition). All such individuals developed high level of ACE2 blocking antibodies following immunization.

**Figure 3:**
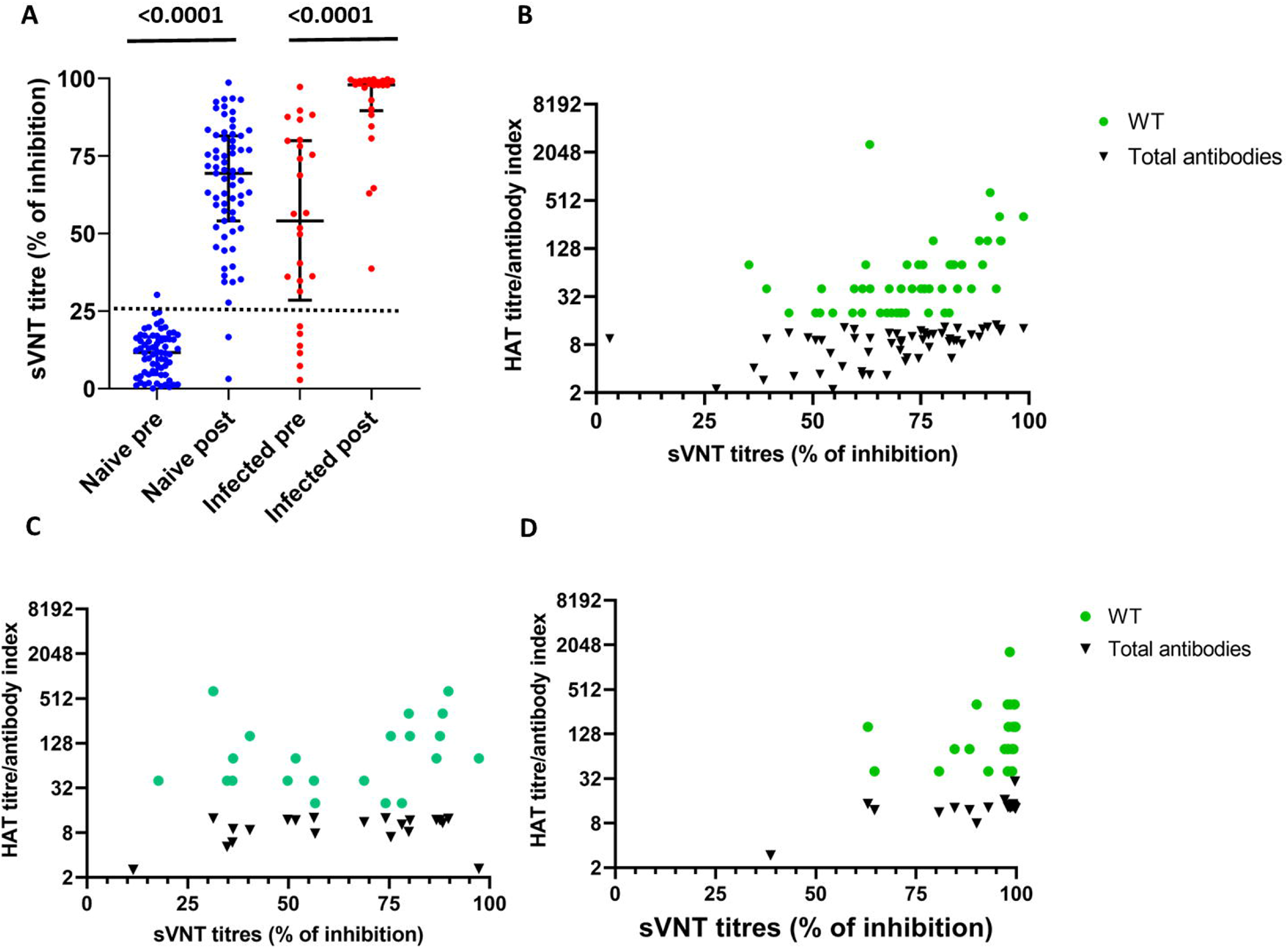
Surrogate SARS-CoV-2 neutralising antibody assay (sVNT) in individuals who were naïve and previously infected following a single dose of the AZD1222 vaccine. The sVNT titres (% of inhibition) were measured in naïve individuals (blue) and previously infected individuals (red) 28-32 days following the vaccine (A). The sVNT titres were correlated with the HAT titres for the WT virus (Spearman’s R=0.71, p<0.0001), and the SARS-CoV-2 specific total antibodies (Spearman’s R=0.54, p<0.0001) (B). The sVNT titres were correlated with the HAT titres for the WT virus (Spearman’s R=0.64, p=0.0005) and the SARS-CoV-2 specific total antibodies (Spearman’s R=0.56, p=0.003) in previously infected individuals at baseline (C), and 28-32 days following a single dose of the vaccine for the WT virus (Spearman’s R=0.47, p=0.01and the SARS-CoV-2 specific total antibodies (Spearman’s R=0.25, p=0.21) (D). The black dotted line indicates the positive cut-off for ACE2 blocking antibodies in (A) and for the HAT in (B,C and D).

The sVNT titres (ACE2 blocking antibodies) correlated significantly with the HAT titres for the WT virus (Spearman’s R=0.71, p<0.0001)and with the SARS-CoV-2 specific total antibodies (Spearman’s R=0.53, p<0.0001) in those who were previously naïve, suggesting that the ACE2 blocking antibodies and RBD antibodies increased similarly following the vaccine in these individuals (Figure 3B). At the time of recruitment of those who were previously infected, the sVNT titres correlated significantly with the HAT titres for the WT virus (Spearman’s R=0.63, p=0.005), and with the SARS-CoV-2 specific total antibodies (Spearman’s R=0.56, p=0.003) (Figure 3C). In these individuals, the sVNT titres significantly correlated with the HAT titres for the WT virus (Spearman’s R=0.47, p=0.01), following vaccination but not with the total antibodies post vaccination (Figure 3D).

Of the 69 naïve individuals, 2 individuals did not develop ACE2 blocking antibodies or antibodies to the RBD following vaccination, while 13 of those who did not appear to have detectable antibodies to the RBD by HAT, had ACE2 blocking antibodies. However, the ACE blocking antibody titres were significantly less (p<0.0001) in those who were negative by the HAT for antibodies (median 45, IQR 34.3 to 56.8 % of inhibition), compared to those who were positive by the HAT assay (median 74.7, IQR 63.2 to 83.3 % of inhibition).

In the previously infected individuals, the median HAT titres increased from a median of 40 (IQR 20 to 160) to a median of 120 (IQR70 to 200) following a single dose of the vaccine. Interestingly, the increase was more for ACE2 blocking antibodies in previously infected individuals, which increased from 54.1% to 97.9%, suggesting that the increase of antibodies to the RBD is likely to be shifted towards the ACE2 blocking antibodies in those who were previously infected.

### *Ex vivo* T cell responses to overlapping peptides of the spike protein

We investigated the *ex vivo* IFNγ ELISpot responses in 76 individuals, to two overlapping pools representing the spike protein, S1 (peptide 1 to 130) and S2 (peptides 131 to 253). Of the 76 individuals, 4 individuals were previously infected with SARS-CoV-2. Of SARS-CoV-2 naïve individuals, only 2/72 had *ex vivo* T cell responses to the S1 pool of peptide pre-vaccination, possibly due to cross reactivity with other seasonal coronaviruses. None of the naïve individuals had any responses to the S2 pool of peptides pre-vaccination. The *ex vivo* IFNγ ELISpot responses to both S1 and S2 significantly increased (p<0.0001) (Figure 4A). The responses to the S1 pool of peptides representing the early (peptide 1 to 130) region of the spike protein (median 397.5, IQR 165.0 to 702.5 SFU/1 million PBMCs) compared to the S2 pool (peptide 131 to 256) of overlapping peptides (median 155, IQR 75 to 417.5, SFU/1 million PBMCs). There were no significant differences to either S1 (p=0.57) or S2 (p=0.06), between the different age groups (Figure 4B). A *ex vivo* ELISpot response of the mean±2 SD of the background responses was considered as a positive response. 46/72 (63.9%) of individuals had responses to the S1 pool of peptides and 23/72 (31.9%) had a positive response to the S2 pool of peptides.

**Figure 4:**
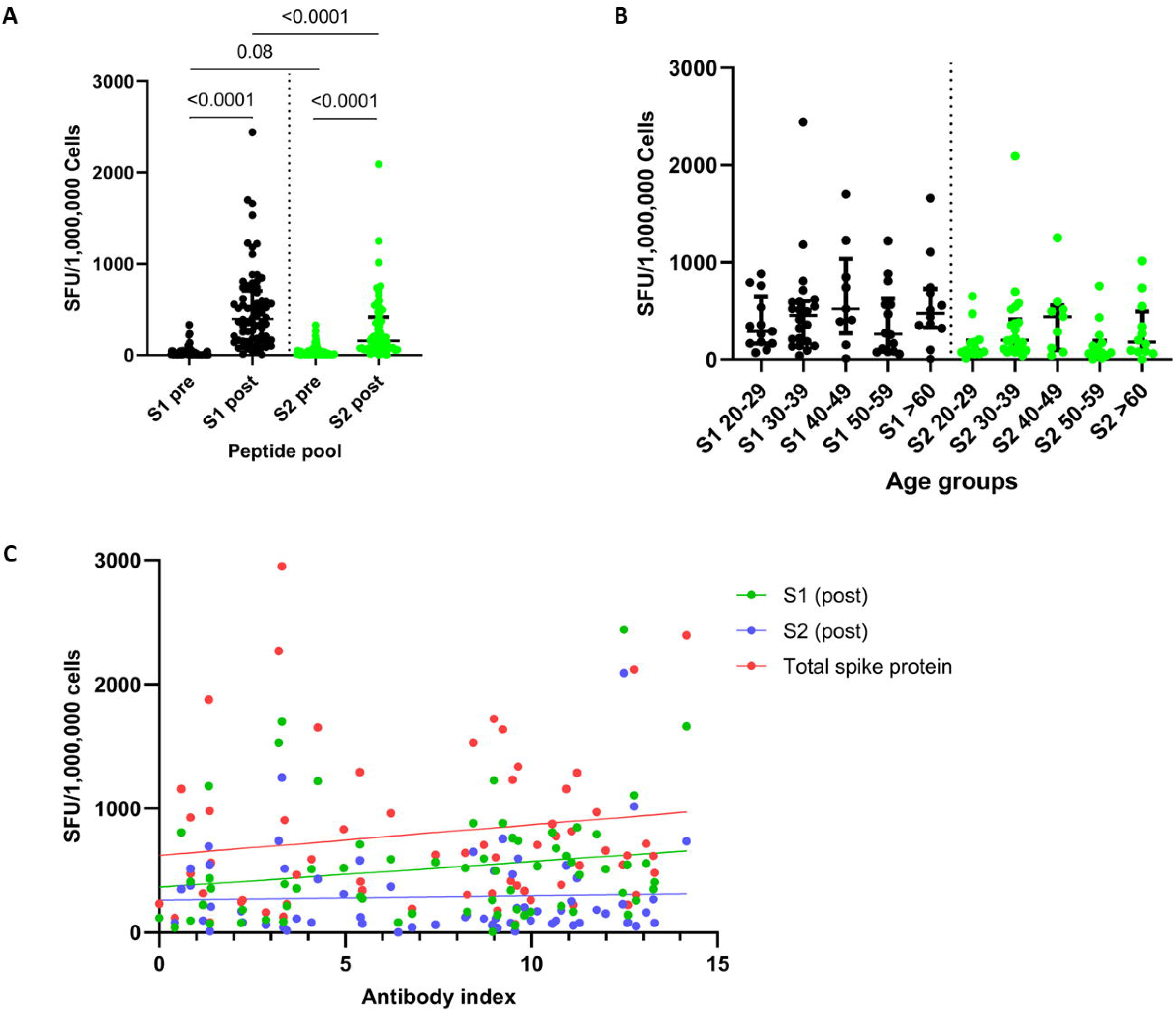
Ex vivo IFNγ ELISpot responses in individuals at baseline and 28 to 32 days following a single dose of the AZD1222/Covishield vaccine. *Ex vivo* IFNγ ELISpot responses were measured to two pools representing the spike protein (S1 and S2) at the baseline (pre) and 28 to 32 days following the vaccine in total naïve individuals (A), and in different age subgroups of naïve individuals (B). The association of the *ex vivo* IFNγ ELISpot responses to the two pools of the spike protein (S1 and S2) and the total responses to overlapping peptides of the spike protein did not correlate with the total antibody responses (C).

The *ex vivo* IFNγ ELISpot responses to S1, S2 or the total S protein did not correlate with the total antibody titres specific for SARS-CoV-2 (Figure 4C). The *ex vivo* ELISpot responses also did not correlate with the HAT titres for the WT (Spearman’s R=-0.08, p=0.48) or with the % of inhibition (ACE-Abs) given by the sVNT assay (Spearman’s R=0.02, p=0.86).. There were no significant differences in the HAT titres in those who responded to S1 (p=0.34) and S2 pool (p=0.86) of peptides compared to those who did not respond to these peptides. There were also no significant differences in the ACE2 blocking antibodies (% of inhibition) in those who responded to S1 (p=0.66) and S2 pools (p=0.42) of peptides, compared to those who had no responses. One of the two individuals who had no antibody responses to the vaccine also did not generate any T cell responses, while the other person did have detectable T cell responses.

Of four individuals who were previously infected with SARS-CoV-2, three had a very low frequency of *ex vivo* IFNγ ELISpot responses pre-vaccination to both S1 (median 47.5, IQR 33.75 to 428.5 SFU/1 million PBMCs) and S2 (median 152.5, IQR 108.8 to 192.5 SFU/1 million PBMCs). The fourth individual was later found to have an acute infection. Following immunization, the frequency of ex vivo T cell responses increased several fold in those with past COVID-19.

## Discussion

In this study we have investigated antibody and *ex vivo* T cell responses to a single dose of the AZD1222 vaccine 28 to 32 days following immunization in previously naïve and infected HCWs. Our results show that 92.9% previously naive individuals seroconverted to a single dose of the vaccine, irrespective of age and gender. A single dose of the vaccine was found to induce similar magnitude antibody and T cell responses in those who were <60 years of age and >60 years of age, although the seroconversion rates were lower in >60-year-olds compared to younger individuals. In naïve individuals, a single dose appeared to induce a higher proportion of ACE2 blocking antibodies than following natural infection. Our previous data in the Sri Lankan individuals with natural COVID-19 infection, showed that although all individuals with moderate to severe illness had ACE2-Abs, assessed by the sVNT assay, 23/69 (33.3%) with mild illness did not have a response above the positive cut-off value (>25% of inhibition)^23^. In contrast, only 2/69 (2.9%) of previously naïve individuals failed to have a positive NAb following a single dose of the vaccine. Similar results were seen with the HAT assay following natural infection and immunization. For instance, only 33/66 (50%) with those with asymptomatic/ mild illness had a positive antibody response by the HAT assay for the WT virus at the end of 4 weeks (under review), whereas 78.2% had a positive response to the RBD antibodies by the HAT assay following a single dose of vaccine. Therefore, a single dose of the AZD1222 vaccine appears to induce a robust SARS-CoV-2 antibody response targeting the RBD of the virus, which is thought to associate with protection.

Individuals who have recently recovered from natural COVID-19 infection were shown to have robust CD4+ and CD8+ T cell responses to many of the viral proteins, which were of a higher magnitude and breadth in those who had experienced severe illness ^24^. 18% to 32% of individuals were found to recognize different regions of the spike protein^24^. The T cell response frequencies were shown to be between 67% to 87% in individuals with mild illness in the convalescent phase or in exposed family members ^25^. We found that 63.9% of individuals showed IFNγ *ex vivo* T cell responses to the S1 pool of overlapping peptides, following a single dose of the vaccine, which is comparable to what was seen following natural infection. The *ex vivo* ELISpot responses observed in our cohort following a single dose of the AZD1222 were slightly higher (median 397.5 for S1 pool and median 155 for S2 pool, SFU/1 million PBMCs) compared to the *ex vivo* ELISpot responses following a single dose of the BNT162b2 (Pfizer BioNTech) vaccine (median 58, SFU/1 million PBMCs)^10^. However, these variations could be due to assay variation between the laboratories, rather than a difference in the T cell responses induced by the two vaccines.

Although the World Health Organization and many other policy makers have recommended that those who have previously been infected with SARS-CoV-2 should obtain the vaccine^26^, many such individuals have been hesitant. However, a single dose of the AZD1222 vaccine in previously exposed individuals not only significantly increased their ACE2 blocking antibodies, but also significantly increased the RBD antibodies for the variants such the B.1.1.7 and B.1.351. The ACE2 blocking titres measured by the sVNT increased from a median of 54.1 to 97.9 % of inhibition, in these individuals. Since a single dose resulted in a substantial increase in the responses in previously infected individuals and also the antibody responses to variants also significantly increased, it would be important to consider if a single dose of the vaccine would provide sufficient immunity in such individuals. In settings where the P1 or B.1.351 variants are causing severe disease even in individuals previously infected with the original SARS-CoV-2, our results suggest a single dose of vaccine based on the original sequence may still induce a significant increase in antibodies crossreactive with the variants – perhaps sufficient to ameliorate disease.

Two (2/69%) naïve individuals did not have any responses to the vaccine (antibodies to RBD and ACE2 blocking antibodies), while one of these individuals had T cell responses. However, of the whole cohort of individuals 7.1% (43/607), had no detectable antibodies by the Wantai total antibody ELISA, which detects IgM, IgA and IgG to the RBD, while 21.8% were negative by HAT. Except for the seroconversion rates being lower in individuals >60 years of age (7/43 who didn’t seroconvert), comorbidities did not affect seroconversion. It would be important to find out if these individuals who had a poor serological response to the vaccine, would be more susceptible to infection in future in prospective studies.

In summary, a single dose of the AZD1222 vaccine induced high levels of antibodies to the RBD and ACE2 blocking antibodies, in previously naïve individuals, which was greater than immune responses in those who experience a mild or asymptomatic natural infection. The T cell responses were comparable to those following natural infection. In those who previously had COVID-19, a single dose induced very high levels of ACE2 blocking antibodies and antibodies to RBDs of SARS-CoV-2 variants of concern.

## Methods

633 HCWs, who received their first dose of the AZD1222/Covisheild vaccine between the 29^th^ January to 5^th^ of February 2021, were included in the study following informed written consent. Demographic details such as age, gender, comorbid illnesses were recorded. Blood samples were obtained from all individuals to determine the SARS-CoV-2 serostatus at baseline, while T cell study were carried out in only 76 individuals. A second blood sample was obtained between 28 days to 32 days following the first dose to assess SARS-CoV-2 specific antibody and T cell responses. Ethics approval was obtained from the Ethics Review Committee of University of Sri Jayewardenepura. None of the individuals included in this study reported any COVID-19 infection during this one month.

### Detection of total antibodies to SARS-CoV-2

SARS-COV-2 specific total antibody (IgM, IgG and IgA) responses were assessed using WANTAI SARS-CoV-2 Ab ELISA (Beijing Wantai Biological Pharmacy Enterprise, China). This assay was shown to have a sensitivity of 98% ^27^ and was found to be 100% specific in serum samples obtained in 2018, in Sri Lankan individuals. The assay was carried out and results were interpreted according to manufacturers’ instructions.

### Haemagglutination test (HAT) to detect antibodies to the receptor binding domain (RBD)

The HAT was carried out as previously described ^21^. The B.1.1.7 (N501Y) and B.1.351 (N501Y, E484K, K417N) versions of the IH4-RBD reagent were produced as described ^21^, but included the relevant amino acid changes introduced by site directed mutagenesis. These variants were titrated in a control HAT with the monoclonal antibody EY-6A (to a conserved class 4 epitope^21,28^) and found to titrate identically with the original version so 100ng (50ul of 2ug/ml stock solution) was used for developing the HAT. Briefly, red blood cells from an O negative donor were mixed with the IH4-RBD (a nanobody against a conserved glycophorin A epitope on red cells, linked to the RBD of SARS-CoV-2) and incubated for one hour with serum. Phosphate buffered saline was used as a negative control. At the end of the incubation the plate was tilted for 20 seconds and then photographed. The photograph of the plate was read by two independent readers to examine the “teardrop” formation indicative of a negative result. A complete absence of “teardrop” formation was scored as positive, and any flow of “teardrop” was scored as negative. The HAT titration was performed using 11 doubling dilutions of serum from 1:20 to 1:20480, to determine presence of RBD-specific antibodies. The RBD-specific antibody titre for the serum sample was defined by the last well in which the complete absence of “teardrop” formation was observed. RBD-specific antibody titres were also evaluated for the RBD of the B.1.1.7 variant and the B.1.351 variant in 69 individuals, who were seronegative at the baseline and in 26 who had previous infection with SARS-CoV-2.

### Measuring the presence of neutralising antibodies to the SARS-CoV-2 using a surrogate assay

Due to the lack of a BSL-3 facility to assess the presence of neutralizing antibodies, we adopted a recently developed surrogate virus neutralization test (sVNT)^22^, which measures the percentage of inhibition of binding of the RBD of the S protein to recombinant ACE2 (Genscript Biotech, USA). Inhibition percentage ≥ 25% in a sample was considered as positive for ACE2 blocking antibodies. This assay was found to be 100% specific for measuring ACE2 blocking antibodies in the Sri Lankan population^23^.

### *Ex vivo* ELISpot assay

Ex vivo IFNγ ELISpot assays were carried out as previously discussed using freshly isolated peripheral blood mononuclear cells (PBMC) obtained from 76 individuals at the time of recruitment and 28 to 32 days later ^29^. Two pools of overlapping peptides named S1 (peptide 1 to 130 and S2 (peptide 131 to 253) covering the whole spike protein (253 overlapping peptides) were added at a final concentration of 10 µM and incubated overnight as previously described ^24,30^. All peptide sequences were derived from the wild-type consensus and were tested in duplicate. PHA was included as a positive control of cytokine stimulation and media alone was applied to the PBMCs as a negative control. The spots were enumerated using an automated ELISpot reader (AID Germany). Background (PBMCs plus media alone) was subtracted and data expressed as number of spot-forming units (SFU) per 10^6^ PBMCs. A positive response was defined as mean±2 SD of the background responses.

### Statistical analysis

GraphPad Prism version 6 was used for statistical analysis. As the data were not normally distributed, differences in means were compared using the Mann-Whitney U test (two tailed), and the Wilcoxon matched-pairs signed rank test was used when comparing paired data. The Kruskal-Wallis test was used to compare the means of the antibody levels and *ex vivo* ELISpot responses in different age groups. Spearman rank order correlation coefficient was used to evaluate the correlation between variables including the association between SARS-CoV-2 specific T cell responses, age and antibody responses.

## Data Availability

All data is included in the manuscript and figures.

## Acknowledgements

We are grateful to the World Health Organization, UK Medical Research Council and the Foreign and Commonwealth Office for support. T.K.T. is funded by the Townsend-Jeantet Charitable Trust (charity number 1011770) and the EPA Cephalosporin Early Career Researcher Fund. A.T. are funded by the Chinese Academy of Medical Sciences (CAMS) Innovation Fund for Medical Science (CIFMS), China (grant no. 2018-I2M-2-002).

## Data sharing

All data is included in the manuscript and figures. Deidentified participant data can be made available on request.

